# “Multiplex RT-PCR for SARS-CoV-2 variant surveillance in resource-limited settings: an in-house validation study in Cuba”

**DOI:** 10.64898/2026.06.22.26356299

**Authors:** Yulaimy Batista Lozada, Yesenia María García Frómeta, Yaimé González González, Yanin Mokdse Beltran, Iria García de la Rosa, Daniel Gutiérrez Luis, Mónica López de Torner, Ariadne Blanco Alarcón, Sheila Triana Mansito, Ana María Rodríguez Suárez

**Affiliations:** Molecular Biology Department, ImmunoAssay Center; Republic University, Uruguay; Monoclonal Antibodies Department, ImmunoAssay Center; Research and Development Department, ImmunoAssay Center

## Abstract

**Background:** SARS-CoV-2 genomic surveillance is vital for public health, but whole-genome sequencing (WGS) remains costly and inaccessible in many resource-limited settings. We developed and validated a multiplex real-time RT-PCR assay for rapid, economical detection of key mutations associated with variants of interest (VOI) and concern (VOC).

**Methodology:** Two multiplex mixes (M1, M2) targeting eight mutations in the *ORF1a* and *Spike* genes were designed. Analytical validation included sensitivity, specificity, reproducibility, and limit of detection (LoD) using WHO international standards and a respiratory pathogen panel. In parallel, an *in silico* analysis evaluated oligonucleotide efficacy against 10.4 million SARS-CoV-2 genomes from GISAID/NCBI, assessing inclusivity, target-site secondary structure (RNAalifold), and hybridization energy (Primer3Plus).

**Results:** The assay demonstrated 100 % clinical sensitivity among samples with valid RT-PCR results (41/42 samples yielded interpretable results, with one inhibited sample excluded from sensitivity calculation), a LoD of 5.7 log₁₀ IU/mL, and 100 % analytical specificity against 32 non-SARS-CoV-2 respiratory pathogens. Six out of eight oligonucleotide sets showed >96 % inclusivity; two sets exhibited reduced inclusivity (94.03 %, 90.14 %) and structural features potentially affecting binding against emerging variants. The assay enables direct identification of major VOCs (Alpha, Beta, Gamma, Delta, Omicron) and indirect detection of multiple VOIs (P.2, Epsilon, Kappa, Eta, Iota, Lambda).

**Conclusion:** This standardized multiplex assay provides a rapid, sensitive, and low-cost alternative for SARS-CoV-2 variant surveillance in Cuba and similar settings. The integration of experimental and *in silico* validation offers a robust, adaptable framework to sustain diagnostic accuracy amid viral evolution, optimizing the allocation of scarce sequencing resources.

**Author Summary:** Genomic surveillance is a cornerstone of the public health response to SARS-CoV-2, yet whole-genome sequencing capacity remains out of reach for many laboratories in low- and middle-income countries. To bridge this gap, we developed and validated a multiplex real-time RT-PCR assay that detects eight key mutations in the viral genome, enabling the identification of 11 priority lineages. The test costs approximately 5 USD per sample, produces results in a few hours, and uses standard thermocyclers — making it suitable for decentralized deployment. Laboratory validation confirmed high diagnostic performance with no cross-reactivity against 32 other respiratory pathogens. Crucially, we combined this experimental work with a large-scale computational analysis of over 10 million publicly available viral genomes. This *in silico* step allowed us to verify that most of our primer and probe sets remain effective against global viral diversity, and to identify two sets that may require future optimization. Our work provides a practical, sustainable model for variant monitoring in settings with constrained sequencing capacity, as exemplified by Cuba. By integrating robust laboratory validation with ongoing bioinformatic surveillance, this framework optimizes the use of scarce genomic resources and strengthens pandemic preparedness in similar regions worldwide.

**Graphical abstract:** 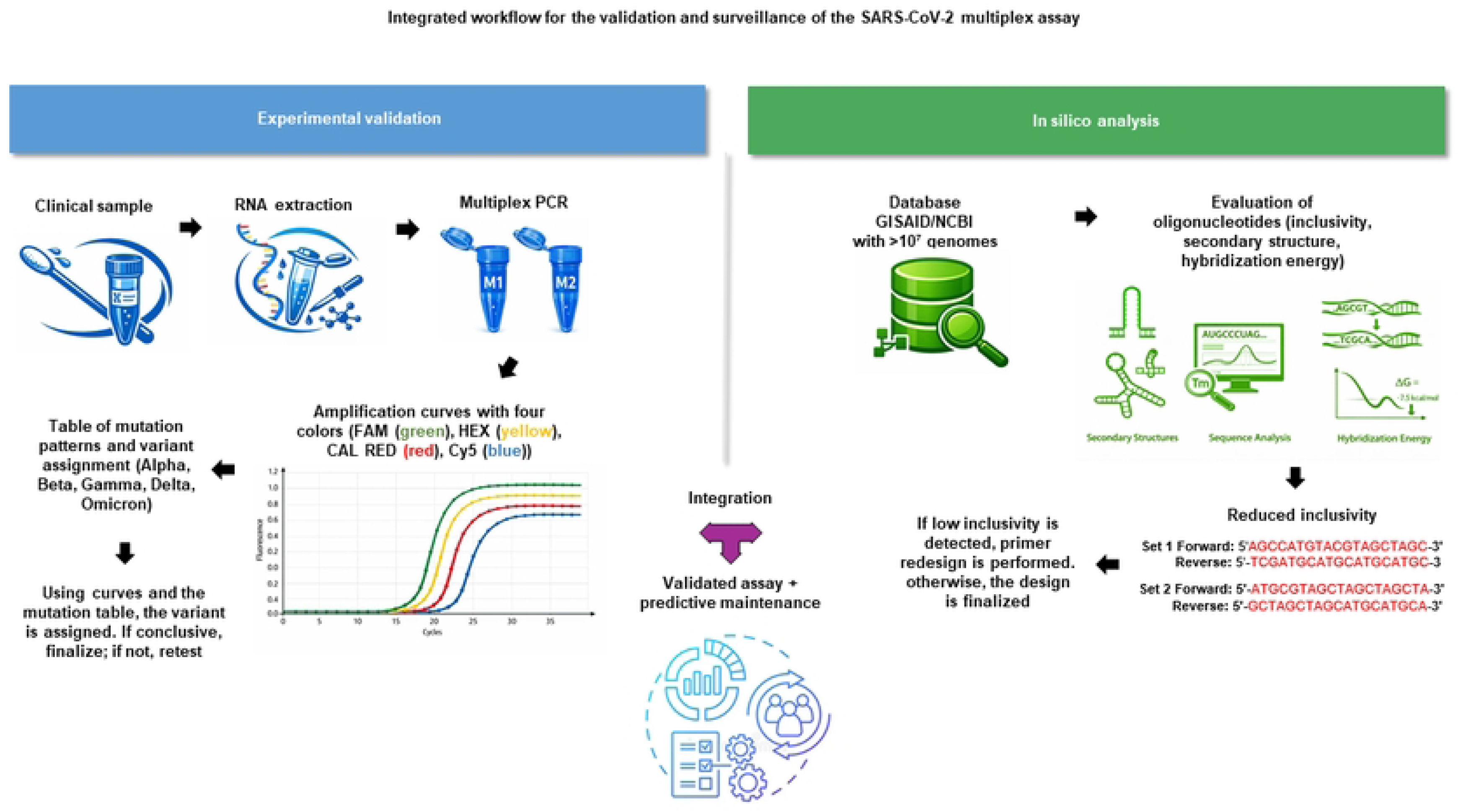

## 1. Introduction

The COVID-19 pandemic has starkly highlighted global inequities in pathogen surveillance capacity [1]. While high-income countries routinely employ whole-genome sequencing (WGS) as the gold standard for tracking SARS-CoV-2 variants, low- and middle-income countries (LMICs) face profound technical and economic barriers [2, 3]. This surveillance gap carries significant public health consequences, as variants of concern (VOCs) like Beta (associated with increased severity) and Delta (characterized by high transmissibility) have had a disproportionate impact in tropical and subtropical regions, undermining vaccine efficacy and increasing mortality rates [4–6]. In Cuba, the national reference laboratory at the Pedro Kouri Institute (IPK) successfully identified multiple imported and community-transmitted variants [7]. However, reliance on centralized WGS created bottlenecks, limiting scalability to peripheral laboratories and delaying timely public health interventions—a challenge echoed across Latin America and the Caribbean [8].

Real-time reverse transcription PCR (RT-PCR) remains the diagnostic cornerstone in LMICs due to its affordability and widespread deployment. Yet, most available assays only confirm the presence of the virus, failing to provide lineage-defining information [9]. In contrast, sequencing, while definitive, demands specialized infrastructure (equipment costs ∼500,000 USD), expensive imported reagents (∼90 USD per sample), and a processing time of 3–7 days, rendering it impractical for routine, high-throughput surveillance in resource-constrained settings [3, 10]. There is therefore an urgent need for accessible, rapid methods to screen for variants [1]. Such tools can effectively triage samples for confirmatory WGS, optimizing the use of scarce sequencing resources and strengthening early warning systems [2].

To address this need, we developed and validated a multiplex real-time RT-PCR assay that simultaneously detects eight key mutations in the SARS-CoV-2 genome, enabling the identification of 11 priority lineages. Our approach integrates rigorous experimental validation with comprehensive *in silico* bioinformatic analysis, drawing on design principles demonstrated for variant screening [11,12]. The assay was designed with core principles of accessibility (approximate cost of 5 USD per test), decentralization (compatibility with standard thermocyclers), and complementarity with existing genomic surveillance networks [13]. This study evaluates the analytical and operational performance of this assay and proposes a scalable model for integrated variant surveillance within LMIC laboratory networks, using Cuba as a case study.

## 2. Materials and Methods

### 2.1 Multiplex assay design and study workflow

#### 2.1.1 Assay design

Based on an analysis of SARS-CoV-2 genomes obtained from GISAID and GenBank, we designed two multiplex mixes (M1 and M2) to detect four mutations each in a single fluorescence channel (Table 1), using TaqMan probes labeled with distinct fluorophores (FAM, HEX, CAL Red 610, Cy5). The assay targets a combination of deletions, single-nucleotide polymorphisms (SNPs), and an insertion, distributed across the ORF1a and Spike genes to maximize coverage of circulating variants. All primers and probes were designed to function with a single annealing temperature (58 °C) and a unified thermal cycling protocol (reverse transcription at 55 °C for 20 min; initial denaturation at 95 °C for 15 s; 42 cycles of denaturation at 95 °C for 15 s, combined annealing and extension at 58 °C for 25 s, with a final acquisition step at 72 °C for 15 s). Table 1 details the target mutations, their genomic location, and the corresponding fluorophore for each channel.

**Table 1.** Design of multiplex mixes for the detection of SARS-CoV-2 variants.

| Mix | Target Mutation | Gene | Fluorophore |
| --- | --- | --- | --- |
| **M1** | ORF1a Δ3675/3677 | ORF1a | FAM |
|  | S Δ241/243 | Spike | HEX |
|  | S Δ69/70 | Spike | CAL Red 610 |
|  | S D614G | Spike | Cy5 |
| **M2** | S Δ156/157 | Spike | FAM |
|  | 214 ins | ORF1a | HEX |
|  | S V1176F | Spike | CAL Red 610 |
|  | S L452R | Spike | Cy5 |

#### 2.1.2 Study workflow

The study was structured into two parallel and complementary pipelines: experimental validation and in silico analysis (Fig. 1), which converge to provide a validated assay with a built-in strategy for continuous monitoring.

**Fig. 1.**
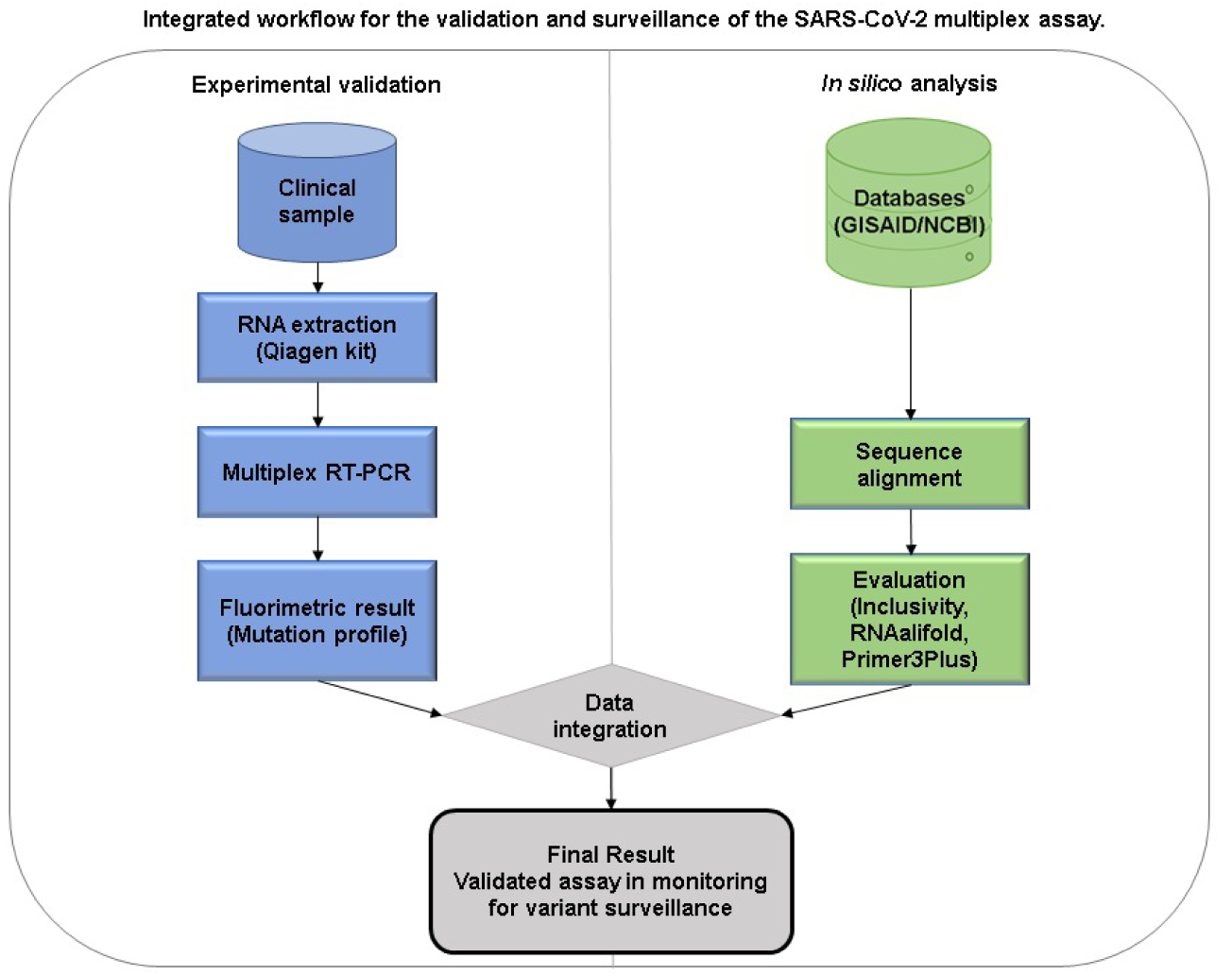
Integrated surveillance model combining experimental workflow and *in silico* monitoring for sustainable SARS-CoV-2 variant tracking.

##### Experimental branch

Nasopharyngeal swab samples from suspected COVID-19 cases were collected and RNA was extracted using the QIAamp Viral RNA Mini Kit (Qiagen). Extracted RNA was then analyzed by the multiplex RT-PCR assay described above on a SLAN-96P Real-Time PCR System. Amplification curves for each fluorescence channel were recorded, and a ΔCt-based algorithm (ΔCt ≤ 5) was used to determine the presence or absence of each target mutation. The resulting mutation profile was cross-referenced against a predefined mutation signature table (derived from public lineage definitions) to assign the corresponding SARS-CoV-2 lineage (e.g., Alpha, Beta, Gamma, Delta, Omicron). Samples with profiles not matching any established signature were flagged for confirmatory whole-genome sequencing (WGS) and further characterization.

##### In silico branch

To assess the long-term robustness of the oligonucleotide sets, a computational analysis was performed using a global dataset of 10,425,874 complete SARS-CoV-2 genomes downloaded from GISAID and NCBI (up to March 2024). For each primer and probe set, we evaluated (i) inclusivity (percentage of genomes with perfect complementarity), (ii) structural stability of the target regions (RNAalifold), and (iii) hybridization energy (ΔG) (Primer3Plus). This analysis identified two oligonucleotide sets with reduced inclusivity (94.03% and 90.14%), which were flagged for priority monitoring in future updates.

The final output (Fig. 1, bottom) integrates the validated experimental assay with a proactive surveillance framework that uses the in silico data to anticipate potential detection failures and guide timely assay updates.

### 2.2 Oligonucleotides and reaction setup

Each mix contained specific forward and reverse primers and dual-labeled hydrolysis probes (FAM/BHQ1, HEX/BHQ1, CAL Red 610/BHQ2, Cy5/BHQ2) for their respective targets. The final composition of each 25 µL multiplex reaction was as follows: 1X Reaction Buffer, 0.35 mM dNTPs, 3.75 U of MMLV reverse transcriptase, 0.46 U of RNasin, and 1.25 U of Taq polymerase. The thermostable polymerase was supplemented with 0.5 mg/mL of Bovine Serum Albumin (BSA) to enhance stability. 10 mM (NH4)_2_SO_4_ was added to optimize amplification efficiency. The final concentrations of each probe ranged from 20 nM to 225 nM depending on the fluorophore used, with primer concentrations set at twice the probe concentration.

### 2.3 Clinical samples, reference materials, and analytical specificity panel

A well-characterized panel of 42 clinical samples, a subset of which were available both as nasopharyngeal swab specimens and as extracted RNA, previously sequenced and lineage-assigned by the national reference laboratory, was used for clinical validation. This panel included 11 Beta (B.1.351), three Alpha (B.1.1.7), eight Epsilon ((B.1.427/429), eight Delta (B.1.617.2), two Omicron (BA.1), two P.1 (Gamma), three D614G (B.1), three B.1.1.732, and two AT.1 variants. Additionally, four samples with known PCR inhibition were included. The limit of detection (LoD) was determined using the WHO International Standard for SARS-CoV-2 RNA (NIBSC 20/146). Analytical specificity was assessed against the NATtrol™ Respiratory Verification 2.0 panel (ZeptoMetrix, Buffalo, NY, USA), which includes 32 viral and bacterial respiratory pathogens (e.g., influenza A/B, RSV, *Streptococcus pneumoniae*, *Mycoplasma pneumoniae*).

### 2.4 RNA extraction and RT-PCR protocol

Viral RNA was extracted from 200 µL of clinical specimen or reference material using the QIAamp Viral RNA Mini Kit (QIAamp Viral RNA Mini Kit, Qiagen, Germany) following the manufacturer’s protocol. Elution was performed in 60 µL of elution buffer. Then, the extracted RNA was used as template to test our assay. Multiplex RT-PCR was performed on a SLAN-96P Real-Time PCR System (Shanghai Hongshi Medical Technology Co., Shanghai, China). The optimized thermal cycling protocol consisted of a reverse transcription step at 55°C for 20 minutes; initial denaturation at 95°C for 15 seconds; followed by 42 cycles of denaturation at 95°C for 15 seconds, combined annealing and extension at 58°C for 25 seconds, and a final signal acquisition step at 72°C for 15 seconds. Fluorescence was measured in the FAM, HEX, CAL Red 610 (or VIC), and Cy5 channels at the end of each cycle.

### 2.5 Lineage assignment algorithm (ΔCt method)

Variant calling from the multiplex assay results was performed using a ΔCt-based algorithm. For each sample and within each multiplex mix (M1 or M2), the difference (ΔCt) was calculated between the highest Ct value obtained (least efficiently amplified target) and the lowest Ct value (most efficiently amplified target). A ΔCt value ≤ 5 was interpreted as specific amplification of the target mutation, and the mutation was considered present. Conversely, a ΔCt value > 5 indicated likely non-specific amplification for the target with the high Ct, and that mutation was considered absent for that sample. The complete mutation profile (presence/absence of each of the eight targets) was then compared to the reference mutation signatures for known variants (Table 2). This algorithm enables direct identification of variants with unique mutation patterns. Importantly, any sample yielding a mutation profile that does not match any of the established patterns is considered a potential indicator of an emerging variant or a recombinant, warranting further investigation by sequencing. This “dynamic detection” approach transforms the assay into an early warning system for novel variants, reinforcing its value in genomic surveillance.

**Table 2.** Expected mutation profiles for major SARS-CoV-2 lineages based on the multiplex RT-PCR amplification patterns.

| Mix | Channel | Mutation | Alpha<br>(B.1.1.7) | Beta<br>(B.1.351) | Gamma<br>(P.1) | Delta<br>(B.1.617.2) | Omicron<br>(BA.1) | Epsilon<br>(B.1.427/B.<br>1.429) |
| --- | --- | --- | --- | --- | --- | --- | --- | --- |
| M1 | FAM | ORF1a<br>Δ3675/3677 | + | + | + | – | + | – |
| | HEX | S Δ241/243 | – | + | – | – | – | – |
| | CAL | S Δ69/70 | + | – | – | – | + | – |
|  | CY5 | S D614G | + | + | + | + | + | + |
| M2 | FAM | S Δ156/157 | – | – | – | + | + | – |
|  | HEX | 214 ins | – | – | – | – | + | – |
|  | CAL | S V1176F | – | – | + | – | – | – |
|  | CY5 | S L452R | – | – | – | + | – | + |

(+) mutation present; (–) mutation absent. Note: For ORF1a Δ3675/3677, S Δ69/70, and S Δ156/157 (wild-type design): absence of amplification indicates mutation present. For S Δ241/243, S D614G, 214 ins, S V1176F, and S L452R (mutant-specific design): amplification indicates mutation present. D614G is present in virtually all post-Wuhan variants and serves as a control for SARS-CoV-2 amplification.

### 2.6 Analytical performance evaluation

#### 2.6.1. Analytical performance

The limit of detection (LoD), defined as the lowest concentration at which 19/20 (95%) replicates were positive, was determined by testing serial dilutions (in triplicate) of the WHO International Standard for SARS-CoV-2 RNA (NIBSC 20/146). Analytical specificity was assessed against the NATtrol™ Respiratory Verification 2.0 panel (ZeptoMetrix, Buffalo, NY, USA), which includes 32 viral and bacterial respiratory pathogens (e.g., influenza A/B, RSV, *Streptococcus pneumoniae*, *Mycoplasma pneumoniae*). Intra-assay and inter-assay reproducibility were evaluated by running triplicates of positive controls (high, medium, low viral loads) in the same run and across three different runs, respectively. Reproducibility was expressed as the coefficient of variation (CV%) of cycle threshold (Ct) values.

#### 2.6.2. Clinical sensitivity and specificity

The capacity of the multiplex assay, combined with the ΔCt-based lineage assignment algorithm (section 2.5), to correctly identify SARS-CoV-2 lineages was evaluated using a well-characterized panel of 42 clinical samples (described in section 2.3), for which whole-genome sequencing (WGS) data and lineage assignments were available from the national reference laboratory. For each sample, RNA was extracted and tested with both M1 and M2 multiplex mixes as per the protocol in section 2.4.

A sample was classified as:

- True Positive (TP) if the assay correctly identified the lineage concordant with WGS.
- False Negative (FN) if the assay failed to identify a lineage that was present according to WGS.
- True Negative (TN) if the assay correctly did not identify a lineage that was absent (i.e., for samples where the targeted mutation profile was not expected).
- False Positive (FP) if the assay assigned a lineage that was not confirmed by WGS.

Samples with PCR inhibition or invalid results were excluded from the calculation of clinical sensitivity and specificity but were reported separately as technical failures. The clinical sensitivity was therefore defined as the proportion of true positives among samples with valid RT-PCR results.

Clinical sensitivity was calculated as:

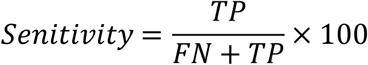

and clinical specificity as:

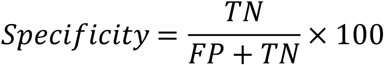

with 95% confidence intervals (CI) computed using the Wilson score method. This analysis allowed us to validate the discriminatory power of the ΔCt threshold in distinguishing specific from non-specific amplification signals in a real-world clinical context.

### 2.7 *In silico* bioinformatic analysis

To evaluate the assay’s robustness and long-term applicability, we conducted a comprehensive in silico bioinformatic analysis using all complete SARS-CoV-2 genomes (n = 10,425,874) available from GISAID and NCBI up to March 2024. After alignment, we assessed each primer and probe set for three key parameters: inclusivity, defined as the proportion of genomes with a perfectly complementary target sequence; structural stability, where RNAalifold [14] was used to predict whether local secondary structures might impede hybridization; and hybridization energy, calculating the Gibbs free energy (ΔG) of binding with Primer3Plus [15] to estimate amplification efficiency. This systematic evaluation enables us to prospectively identify oligonucleotides at risk of failing against emerging variants, thereby guiding timely updates to the assay design.

### 2.8 Statistical analysis

Clinical sensitivity and specificity were calculated with 95 % confidence intervals using Wilson’s score method. Reproducibility was expressed as the CV %. The *in-silico* analysis generated metrics of percentage coverage and energy profiles. All statistical analyses were performed using GraphPad Prism version 9.0 (GraphPad Software, San Diego, CA, USA).

## 3 Results

### 3.1 Analytical validation of the multiplex RT-PCR assay

Following optimization, the multiplex assay demonstrated robust analytical performance suitable for variant surveillance. Among the 42 clinical samples, 41 yielded valid RT-PCR results (97.6 % technical success rate). One sample showed PCR inhibition and was excluded from sensitivity calculations. The clinical sensitivity calculated on the 41 valid samples was 100 % (95 % CI: 91.4–100 %).

The limit of detection (LoD) was determined by testing ten-fold serial dilutions of the WHO International Standard for SARS-CoV-2 RNA (NIBSC 20/146) in triplicate across three independent runs. The LoD, defined as the lowest concentration yielding positive amplification in 19/20 replicates, was established at 5.7 log10 IU/mL (approximately 33.5 Ct on the SLAN-96P system). The assay exhibited 100 % analytical specificity, with no cross-reactivity observed against any of the 32 non-SARS-CoV-2 respiratory pathogens in the NATtrolTM verification panel. Reproducibility was excellent, with intra- and inter-assay coefficients of variation (CV%) for Ct values below 2.5 % and 4.0 %, respectively.

### 3.2 Variant identification profile

The mutation profile targeted by the assay enables a two-tiered identification strategy (Table 2). First, it allows for the direct identification of VOCs and VOIs that were present in the clinical validation panel and for which a unique mutation signature is defined: Alpha (B.1.1.7), Beta (B.1.351), Gamma (P.1), Delta (B.1.617.2), Omicron (BA.1), and Epsilon (B.1.427/429). All these lineages were represented in the validation panel (see Table 3) and were correctly identified by the assay. Second, the assay supports the indirect detection of additional VOIs—including Zeta (P.2), Kappa (B.1.617.1), Eta (B.1.525), Iota (B.1.526), and Lambda (C.37)—which share one or more targeted mutations with the validated lineages. Although these latter variants were not available in the validation panel, their detection is inferred from the mutation profiles and is supported by *in silico* analysis.

**Table 3.** Clinical performance of the multiplex RT-PCR assay against a sequenced panel.

| Lineage (Variant) | No. of samples (WGS-confirmed) | Correctly identified by multiplex assay | Inhibited/Invalid | Concordance (%) |
| --- | --- | --- | --- | --- |
| Beta (B.1.351) | 11 | 11 | 0 | 100 |
| Alpha (B.1.1.7) | 3 | 2 | 1 | 66.7* |
| Epsilon (B.1.429) | 8 | 8 | 0 | 100 |
| Delta (B.1.617.2) | 8 | 8 | 0 | 100 |
| Omicron (BA.1) | 2 | 2 | 0 | 100 |
| P.1 (Gamma) | 2 | 2 | 0 | 100 |
| D614G (B.1) | 3 | 3 | 0 | 100 |
| B.1.1.732 | 3 | 3 | 0 | 100 |
| AT.1 | 2 | 2 | 0 | 100 |
| **Total** | **42** | **41** | **1** | **97.6 % (overall)** |

\*Sensitivity and specificity calculated on valid samples (n=41) were 100%. The 97.6 % represents the proportion of samples yielding interpretable results (technical success rate)

Samples producing profiles consistent with indirect detection require confirmation through epidemiological context or sequencing, thereby enabling efficient triage of samples for resource-intensive whole-genome sequencing. Moreover, any sample that yields a mutation pattern not matching the established profiles is flagged as a potential emerging variant, prompting immediate sequencing and further characterization—a key feature of our dynamic detection approach. Representative examples of the mutation profiles obtained for Alpha and Omicron samples are illustrated in Figs 2 and 3, respectively. These figures show the amplification patterns across the eight targets, highlighting how the unique combination of positive and negative signals allows direct lineage identification as described above.

**Fig. 2.**
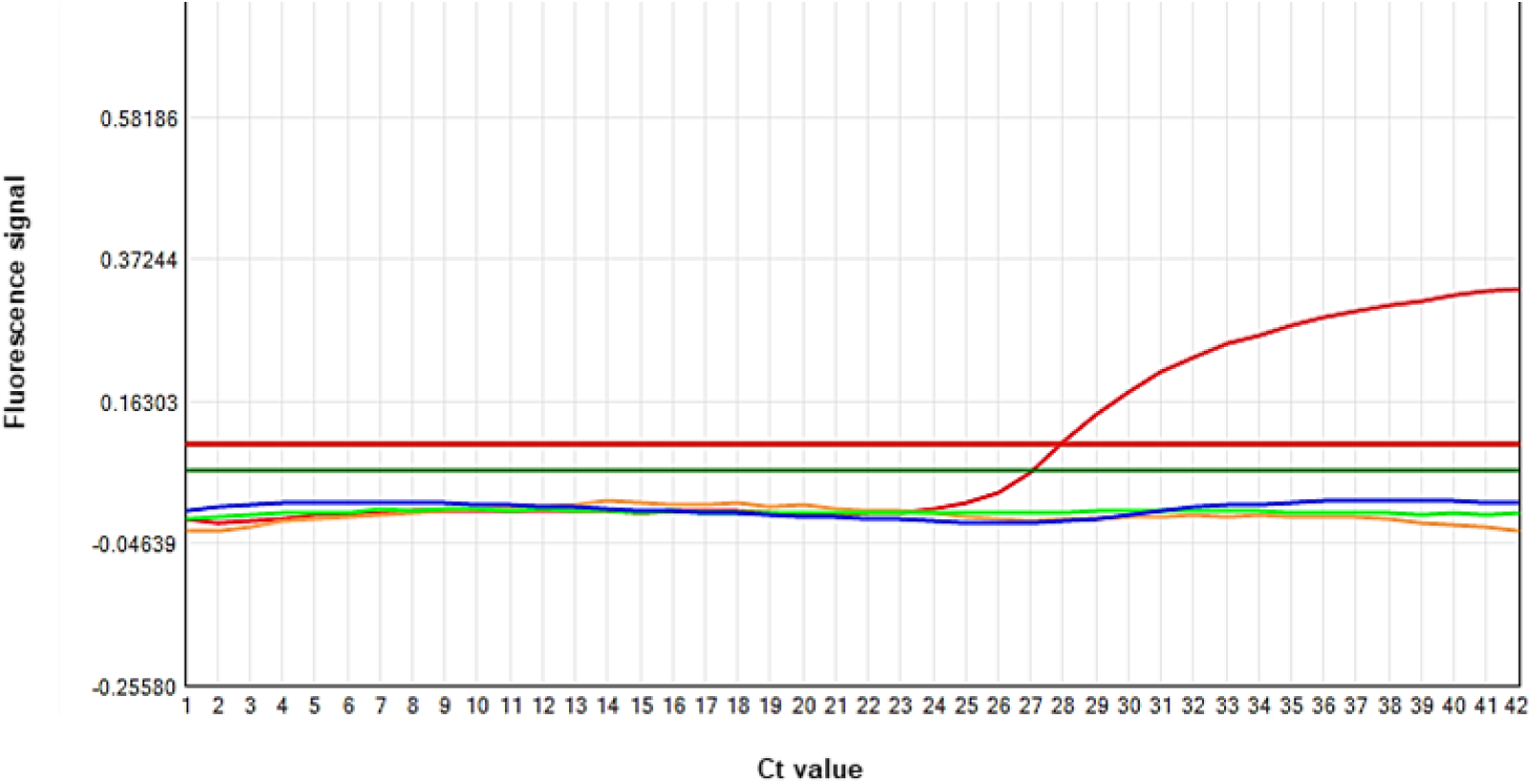
Amplification profile of an Alpha variant (B.1.1.7) sample in multiplex mix M1. Only the Cy5 channel (red curve), targeting the D614G mutation, shows a positive amplification signal. This mutation serves as a control for SARS-CoV-2 positivity and is present in all post-Wuhan variants. The absence of signals in the FAM, HEX, and CAL channels reflects the presence of the characteristic deletions in the Alpha lineage (ORF1a Δ3675/3677, S Δ241/243, and S Δ69/70), which prevent primer or probe binding at those specific targets. The amplification pattern confirms the variant identity while demonstrating the specificity of the assay.

**Fig. 3.**
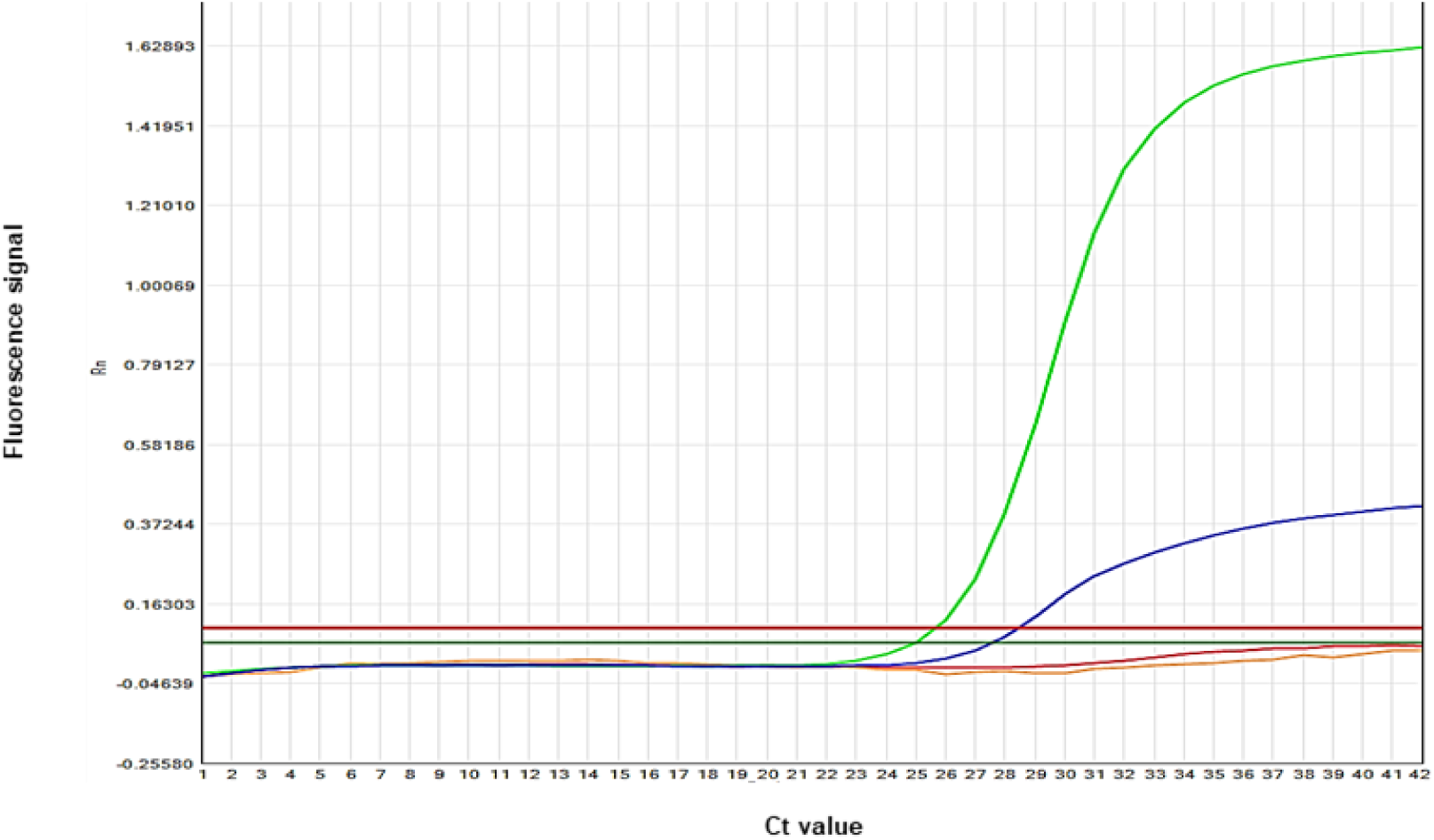
Amplification profile of an Omicron variant (BA.1) sample in multiplex mix M2. Positive signals are observed in the FAM channel (blue curve), corresponding to the S Δ156/157 deletion, and in the HEX channel (green curve), corresponding to the 214 insertion in ORF1a—a characteristic marker of the Omicron BA.1 lineage. The absence of signal in the CAL and Cy5 channels (targeting V1176F and L452R, respectively) is consistent with the expected mutation profile for this variant. This pattern illustrates the assay’s capacity to discriminate Omicron from other variants through its unique combination of targets.

### 3.3 Clinical performance of the ΔCt-based lineage assignment

The clinical accuracy of the multiplex assay was assessed against a panel of 42 sequenced clinical samples comprising 11 Beta (B.1.351), three Alpha (B.1.1.7), eight Epsilon (B.1.429), eight Delta (B.1.617.2), two Omicron (BA.1), two Gamma (P.1), three D614G (B.1), three B.1.1.732, and two AT.1 variants. Of these, 41 samples yielded valid amplification profiles; one Alpha variant sample showed evidence of PCR inhibition (failure to amplify across multiple targets) and was excluded from the primary analysis of valid samples, as no reliable lineage assignment could be made.

Using the ΔCt ≤ 5 threshold to define specific target detection, the assay correctly identified the variant lineage in all 41 valid samples, yielding 41 true positives and zero false positives or false negatives. The single inhibited sample was recorded as an invalid result but not as a false negative, as inhibition was confirmed. Consequently, the clinical sensitivity among samples with valid RT-PCR results was 100 % (41/41; 95 % CI: 91.4–100 %), and the clinical specificity was also 100 % (95 % CI: 91.4–100 %) based on the absence of any misassignments.

The overall concordance with sequencing, including the inhibited sample as non-interpretable, was 97.6% (41/42).

Excluding that sample, the clinical sensitivity and specificity were both 100% (41/41), as detailed in Table 3.

In addition to the lineages listed in Table 3, the validation panel included samples corresponding to B.1 (D614G), B.1.1.732, and AT.1 variants. These were correctly identified as SARS-CoV-2 positive due to detection of the D614G mutation (viral amplification control), but they did not display a unique mutation pattern allowing direct lineage assignment. This finding is consistent with their genomic profiles, which lack the characteristic mutation combinations defined for the variants of interest or concern included in our panel.

Crucially, the ΔCt criterion proved effective in discriminating specific from non-specific amplification signals. In all cases where a mutation was truly absent based on WGS, the corresponding Ct value for that target was either undetected or yielded a ΔCt > 5 relative to the lowest Ct in the mix, preventing false-positive calls. Conversely, for samples with a known mutation profile, all target amplifications exhibited ΔCt values ≤ 5, confirming the robustness of the threshold. These results demonstrate that the multiplex assay, coupled with the ΔCt interpretation algorithm, provides highly accurate and specific lineage identification for circulating SARS-CoV-2 variants, with the only limitation being sample integrity (e.g., inhibition).

### 3.4 *In silico* assessment of oligonucleotide robustness and future utility

To evaluate the potential long-term utility of the assay against viral evolution, a computational analysis was performed by using a global dataset of over ten million SARS-CoV-2 genomes. Inclusivity analysis, defined as the percentage of genomes with perfectly complementary target sequences, revealed that six of the eight oligonucleotide sets maintained inclusivity greater than 96 %, indicating broad coverage of circulating diversity. However, two oligonucleotide sets showed reduced inclusivity (94.03 % and 90.14 %). Secondary structure analysis using RNAalifold indicated a moderate propensity for stable secondary structure formation in the target regions of these two sets. Coupled with less favorable calculated hybridization energy (ΔG), this suggests these sets may be more susceptible to detection failures if additional mutations arise in these genomic regions. These findings validate the current assay’s robustness while clearly identify priority targets for monitoring and potential redesign in future updates, establishing a proactive framework for assay maintenance (Table 4)

**Table 4.** Summary of analytical performance and oligonucleotide coverage.

| Parameter | Result | Interpretation / Comment |
| --- | --- | --- |
| **Clinical Sensitivity** | 100 % (41/41) | High diagnostic performance. |
| **Technical success rate** | 97.6 % (41/42) | One inhibited sample excluded. |
| **Limit of Detection (LoD)** | 5.7 log<sub>10</sub> IU/mL (Ct ~33.5) | Adequate for moderate-to-low viral load samples. |
| **Analytical Specificity** | 100 % (0/32 cross-reactive pathogens) | No interference from common respiratory pathogens. |
| **Reproducibility (Ct CV%)** | Intra-assay: <2.5 %;<br>Inter-assay: <4.0 % | High assay consistency and robustness. |
| **Inclusivity** | Six high-inclusivity oligos sets (>96 %) | Demonstrates broad coverage against global genomic diversity. |
|  | Two sets for priority monitoring (94.03 %, 90.14 %) | Identified via *in silico* analysis for potential susceptibility to drift. |

## 4. Discussion

This study presents the comprehensive validation of a multiplex RT-PCR assay designed to bridge a critical surveillance gap for SARS-CoV-2 variants in resource-limited settings. By integrating rigorous wet-lab validation with a prospective *in silico* bioinformatic framework, we demonstrate a robust, accessible, and sustainable model for variant monitoring. Our work directly addresses the dual challenge of achieving high diagnostic performance while ensuring assay longevity amidst relentless viral evolution [16]. Integrated proposal was presented at Fig 1.

The diagram illustrates the two interconnected components of our proposed surveillance framework. The experimental workflow (left) begins with RNA extraction from clinical samples, followed by multiplex RT-PCR using mixes M1 and M2. The ΔCt algorithm interprets the amplification patterns to assign lineages, and samples with unresolved or unusual profiles are prioritized for confirmatory whole-genome sequencing (WGS). The *in silico* workflow (right) operates in parallel: periodically, all available SARS-CoV-2 genomes from public databases (GISAID/NCBI) are analyzed to assess oligonucleotide coverage, target-site secondary structure, and hybridization energy. This identifies primer/probe sets at risk of failure due to viral evolution, guiding rational redesign. The bidirectional arrows between the two workflows represent the continuous feedback loop: experimental findings inform *in silico* evaluations, and computational predictions prompt updates to the assay, ensuring long-term reliability. This “predictive maintenance” approach transforms reactive assay replacement into a proactive strategy, safeguarding genomic surveillance investments in resource-limited settings.

### 4.1 Addressing an urgent surveillance gap: the rationale for targeted genotyping

The COVID-19 pandemic has starkly exposed global inequities in genomic surveillance capacity, where reliance on centralized whole-genome sequencing (WGS) creates bottlenecks in low- and middle-income countries (LMICs) [17,18]. This deficit is particularly consequential given the rapid emergence of VOCs that can alter transmissibility, disease severity, and vaccine efficacy [19]. In this context, the need for rapid, decentralized screening tools is unequivocal. Our assay, with an approximate cost of 5 USD per test and a turnaround time of a few hours, provides a pragmatic solution. This model of “genomic surveillance triage” — where high-throughput PCR screening prioritizes samples for confirmatory WGS — has been advocated as an effective strategy to optimize scarce sequencing resources in resource-constrained settings [20,21]. The approach is strongly supported by recent literature that positions RT-PCR genotyping as a complementary, high-throughput pillar of integrated surveillance frameworks [22].

### 4.2 Analytical performance: benchmarking against established standards

The proposed assay achieved a clinical sensitivity of 100 % among samples with valid RT-PCR results, with a technical success rate of 97.6 % (one inhibited sample excluded) and 100 % analytical specificity against a panel of 32 respiratory pathogens. This performance is congruent with studies validating similar laboratory-developed and commercial multiplex assays, which report sensitivities ranging from 97.89 % to 100 % [23,24]. The limit of detection (5.7 log₁₀ IU/mL) is suitable for community surveillance where moderate viral loads are prevalent and aligns with previously reported thresholds for comparable RT-PCR methods [23,25]. Crucially, the high reproducibility (coefficient of variation <4.0% for cycle threshold values) confirms operational robustness in peripheral laboratories [26]. This performance profile validates that locally developed assays can meet the stringent standards required for public health decision-making, offering a cost-effective alternative to proprietary kits without compromising reliability.

### 4.3 Clinical accuracy and the value of ΔCt-based lineage assignment

The clinical validation of our ΔCt-based lineage assignment algorithm demonstrated perfect sensitivity and specificity (100%) among samples with valid RT-PCR results, underscoring the robustness of the multiplex assay for variant surveillance. This performance aligns with other reported multiplex genotyping assays; for instance, Wang *et al.* [22] described a similar approach achieving >98% concordance with sequencing, while Vogels et al. [9] highlighted the importance of threshold-based discrimination to avoid false positives in degenerate targets. The use of ΔCt for variant inference has been previously explored in large-scale surveillance studies;

Bordoy et al. [38] successfully employed ΔCt between the N-gene and RdRp targets to monitor the transition of SARS-CoV-2 variants (Alpha, Delta, Omicron) in over 15,000 clinical samples, achieving high predictive accuracy (sensitivity 91–98%, specificity 90–97%) against WGS-confirmed lineages. This demonstrates that ΔCt-based approaches, whether applied to gene targets (as in Bordoy et al.) or mutation-specific probes (as in our assay), constitute a valuable and scalable strategy for variant tracking in resource-limited settings.

The single inhibited sample encountered in our panel reflects a well-recognized challenge in clinical RT-PCR, particularly with nasopharyngeal swabs, and reinforces the need for including internal amplification controls in routine surveillance workflows [39]. A key strength of our method is the use of ΔCt as an internal quality metric to distinguish genuine mutation-specific signals from background or non-specific amplification. This approach is conceptually similar to the “allelic discrimination” employed in some commercial genotyping kits [40] but adapted for multiplex detection of deletions and SNPs. By requiring a ΔCt ≤ 5 for positivity, we effectively minimized the risk of false assignments due to primer–probe mismatches or low-level cross-reactivity, a concern that has been raised for assays targeting rapidly evolving viruses [33]. The 100 % specificity observed in our panel provides empirical validation of this threshold.

Our assay not only correctly identified the Omicron (BA.1) variant but also demonstrated its ability to detect a broader spectrum of relevant mutations. A notable finding was the detection of the ORF1a Δ3675/3677 mutation in the Omicron sample. This triple-amino-acid deletion in the nsp6 domain of the ORF1ab polyprotein is a shared genomic marker present in the earliest Omicron lineages (BA.1, BA.2, BA.3) as well as in other variants of concern such as Alpha, Beta, and Gamma [41,42]. Although its precise biological function is still under investigation, the literature suggests that this deletion may contribute to increased transmissibility or immune evasion by enhancing nsp6-mediated antagonism of type-I interferon signaling [43]. The confirmation of this deletion in our Omicron sample supports the notion that it is a recurrent mutational hotspot rather than a lineage-specific trait. The ability of our multiplex assay to detect such a cross-lineage molecular signature further consolidates its value as a robust tool for epidemiological surveillance.

Looking forward, the integration of such ΔCt-based algorithms with routine *in silico* monitoring (as in section 3.4) creates a resilient surveillance framework: the experimental cut-off can be periodically re-evaluated against emerging variants to ensure sustained accuracy [44]. This dual approach—computational prediction of oligonucleotide efficacy coupled with empirical ΔCt validation—offers a model for other LMICs seeking to deploy cost-effective genotyping tools without compromising reliability.

### 4.4 The integrated *in silico* framework: a proactive strategy for assay longevity

The principal innovation of our work relies on the systematic coupling of experimental validation with prospective computational analysis. While endpoint validation confirms present utility, it provides no safeguard against future viral evolution. It is well documented that primer-template mismatches arising from mutations can lead to reduced sensitivity or complete assay failure, as witnessed with the S-gene target failure in several commercial kits during the emergence of the Alpha variant [27,28]. To preempt such obsolescence, we evaluated our oligonucleotides against a global dataset of >10 million SARS-CoV-2 genomes.

This analysis transcends simple inclusivity metrics. By employing RNAalifold to predict target-site secondary structures and calculating hybridization energies (ΔG), we identified two oligonucleotide sets with marginally reduced inclusivity (94.03 %, 90.14 %) and a higher propensity for structurally stable target regions that may impede efficient primer/probe binding [29,30]. This provides a data-driven, early-warning system that can guide timely assay updates. Such a proactive, bioinformatics-guided approach is increasingly advocated for the design of durable diagnostic tools, emphasizing the selection of conserved genomic regions and continuous sequence surveillance [31,32]. Our framework thus transforms assay maintenance from a reactive exercise into a predictive one, safeguarding public health investments and ensuring the sustained relevance of the surveillance tool.

### 4.5 Dynamic detection: an early warning system for emerging variants

Beyond its ability to classify known variants, our assay incorporates a dynamic detection feature: any sample yielding a mutation profile that does not match the established patterns in Table 2 is flagged as a potential emerging variant. This is a critical innovation, as it transforms the assay from a mere classification tool into an active surveillance instrument. In practice, such unexpected profiles—for instance, a combination of mutations not previously seen together—would trigger immediate reflex sequencing, enabling early identification of novel variants or recombinants. This concept aligns with the recommendations of Grubaugh *et al.* [31] and others who advocate for flexible, multi-tiered surveillance systems that combine high-throughput screening with confirmatory sequencing. By design, this assay thus contributes not only to monitoring known threats but also to detecting the unexpected, reinforcing pandemic preparedness.

### 4.6 Implications for sustainable pathogen surveillance in LMICs

The model established here — local design, experimental validation, and computational sustainability checks — has implications that extend well beyond SARS-CoV-2. It offers a blueprint for developing affordable multiplex tools for other epidemic-prone and neglected diseases in LMICs [33]. The core principles are transferable: to target conserved or signature genomic regions, to design for compatibility with widely available instrumentation, and to implement routine *in silico* reevaluation against emerging sequence data. This pathway can accelerate the development of integrated, multi-pathogen surveillance panels, a critical need for strengthening health security in tropical and subtropical regions where co-circulation of respiratory viruses and arboviruses is common [34,35]. Our work exemplifies how bioinformatics technologies are fundamental not only to studying viral evolution but also to building resilient diagnostic systems that can adapt to changing pathogen landscapes [36].

### 4.7 Limitations, context, and future directions

We acknowledge the intrinsic limitations of a mutation-specific assay. It is a tool for screening known variants, not for discovering novel ones; thus, it functions as an essential complement to, not a replacement for, broad-based WGS [17,21]. Its effectiveness is contingent on the continued epidemiological relevance of the chosen mutation panel, which requires periodic re-evaluation.

These limitations, however, define clear and actionable future directions. First, the *in silico* data on oligonucleotides with reduced inclusivity guide the rational redesign of primer/probe sets, potentially employing mismatch-tolerant designs or shifting targets to more conserved adjacent regions [37]. Second, the modular nature of the assay facilitates the expansion of the mutation panel to include hallmark mutations of emerging Omicron sub-lineages and other VOIs. Finally, the entire workflow is a candidate for adaptation into a multiplex platform for other respiratory pathogens of public health importance in Cuba and similar settings, such as influenza, respiratory syncytial virus, and tuberculosis [23]

## 5. Conclusions

The multiplex RT-PCR assay developed and validated in this study represents a practical, scalable, and economically accessible solution for SARS-CoV-2 variant surveillance in resource-limited settings. By achieving 100% clinical sensitivity (among valid samples) and a 97.6% technical success rate and specificity (100 %) comparable to commercial kits at a fraction of the cost (∼5 USD per test), our assay addresses a critical gap in the global genomic surveillance architecture [1,17]. More importantly, the integration of experimental validation with a prospective *in silico* sustainability framework introduces a paradigm shift: from reactive assay replacement to predictive maintenance. This approach, enabled by open-access bioinformatic tools [14, 15], allows laboratories in LMICs to monitor oligonucleotide performance against an ever-expanding viral sequence space and to rationally update their diagnostic panels [16, 29].

Beyond SARS-CoV-2, the modular design and the underlying principles — local design, compatibility with standard instrumentation, and iterative computational re-evaluation — are readily transferable to other emerging and re-emerging pathogens of public health importance in tropical and subtropical regions [18, 34]. In the Cuban context, this work establishes a precedent for decentralized genomic intelligence and strengthens the country’s preparedness for future epidemic threats [7, 8]. We therefore contend that similar LMIC-driven initiatives, when supported by equitable open-access policies and North–South collaborative frameworks, can substantially democratize pathogen surveillance and contribute to global health security [3, 35].

## Data Availability

The authors confirm that all data underlying the findings are fully available without restriction. The minimal data set, including cycle threshold (Ct) values, mutation profiles, and lineage assignments for the 42 clinical samples, is provided in the manuscript (Table 3 and Supporting Information). The primer and probe sequences generated during this study are available from the corresponding author upon reasonable request. The SARS?CoV?2 genome sequences used for the in silico analysis are publicly available from the GISAID database (https://gisaid.org) and the NCBI GenBank repository (https://www.ncbi.nlm.nih.gov/genbank/). Accession numbers or GISAID identifiers for the specific genomes analysed are available upon request due to the large volume of data (over 10 million genomes). No additional datasets were generated or analysed during the current study.

## References

1. Duarte JG, de Almeida Campos AC, da Silva TM, et al. The inequality of COVID-19 genomic surveillance. PLoS Negl Trop Dis. 2023;17(10):e0011676. doi:10.1371/journal.pntd.0011676

2. Gangavarapu K, Latif AA, Mullen JL, et al. Outbreak.info genomic reports: scalable and dynamic surveillance of SARS-CoV-2 variants and mutations. Nat Methods. 2023;20:512–522. doi:10.1038/s41592-023-01770-8

3. Gardy JL, Loman NJ. Towards a genomics-informed, real-time, global pathogen surveillance system. Nat Rev Genet. 2018;19(1):9–20. doi:10.1038/nrg.2017.88

4. Tang CY, Boftsi M, Staudt L, et al. SARS-CoV-2 and Dengue virus co-infection: A systematic review of reported cases. PLoS Negl Trop Dis. 2022;16(9):e0010645. doi:10.1371/journal.pntd.0010645

5. Tregoning JS, Flight KE, Higham SL, et al. Progress of the COVID-19 vaccine effort: viruses, vaccines and variants versus efficacy, effectiveness and escape. Nat Rev Immunol. 2021;21:626–636. doi:10.1038/s41577-021-00592-1

6. Abdool Karim SS, de Oliveira T. New SARS-CoV-2 variants — clinical, public health, and vaccine implications. N Engl J Med. 2021;384:1866–1868. doi:10.1056/NEJMc2100362

7. González MA, Pérez LJ, Corrales I, et al. First report of SARS-CoV-2 variant B.1.1.7 (Alpha) in Cuba. Infect Genet Evol. 2021;94:105000. doi:10.1016/j.meegid.2021.105000

8. Sah R, Shrestha S, Mehta R, et al. SARS-CoV-2 variant B.1.1.7 (Alpha) and B.1.351 (Beta) in Latin America and the Caribbean countries: A need for more genomic surveillance. Int J Health Plann Manage. 2022;37(1):297–301. doi:10.1002/hpm.3325

9. Vogels CBF, Brito AF, Wyllie AL, et al. Analytical sensitivity and efficiency comparisons of SARS-CoV-2 RT–qPCR primer–probe sets. Nat Microbiol. 2020;5:1299–1305. doi:10.1038/s41564-020-0761-6

10. de Oliveira Martins L, Sanabani SS, Riediger IN, et al. Challenges for SARS-CoV-2 genome sequencing in a low and middle-income country. J Braz Soc Trop Med. 2021;54:e0835. doi:10.1590/0037-8682-0835-2020

11. Wang H, Miller JA, Verghese M, et al. Multiplex SARS-CoV-2 genotyping reverse transcriptase PCR for population-level variant screening and epidemiologic surveillance. J Clin Microbiol. 2021;59(8):e00859–21. doi:10.1128/JCM.00859-21

12. Freed NE, Vlková M, Faisal MB, Silander OK. Rapid and inexpensive whole-genome sequencing of SARS-CoV-2 using 1200 bp tiled amplicons and Oxford Nanopore Rapid Barcoding. Biol Methods Protoc. 2020;5(1):bpaa014. doi:10.1093/biomethods/bpaa014

13. Tegally H, San JE, Cotten M, et al. The evolving SARS-CoV-2 epidemic in Africa: insights from rapidly expanding genomic surveillance. Science. 2022;378(6615):eabq5358. doi:10.1126/science.abq5358

14. Bernhart SH, Hofacker IL, Will S, et al. RNAalifold: improved consensus structure prediction for RNA alignments. BMC Bioinformatics. 2008;9:474. doi:10.1186/1471-2105-9-474

15. Untergasser A, Nijveen H, Rao X, et al. Primer3Plus, an enhanced web interface to Primer3. Nucleic Acids Res. 2007;35(Web Server issue):W71–W74. doi:10.1093/nar/gkm306

16. Sanjuán R, Domingo-Calap P. Mechanisms of viral mutation. Cell Mol Life Sci. 2016;73(23):4433–4448. doi:10.1007/s00018-016-2299-6

17. Inzaule SC, Tessema SK, Kebede Y, et al. Genomic-informed pathogen surveillance in resource-limited countries. Lancet Microbe. 2021;2(11):e577–e580. doi:10.1016/S2666-5247(21)00260-6

18. Brito AF, Semenova E, Dudas G, et al. Global disparities in SARS-CoV-2 genomic surveillance. medRxiv [Preprint]. 2021. doi:10.1101/2021.08.21.21262393

19. Graham MS, Sudre CH, May A, et al. Changes in symptomatology, reinfection, and transmissibility associated with the SARS-CoV-2 variant B.1.1.7: an ecological study. Lancet Public Health. 2021;6(5):e335–e345. doi:10.1016/S2468-2667(21)00055-4

20. Wilkinson E, Giovanetti M, Tegally H, et al. A year of genomic surveillance reveals how the SARS-CoV-2 pandemic unfolded in Africa. Science. 2021;374(6566):423–431. doi:10.1126/science.abj4336

21. Viana R, Moyo S, Amoako DG, et al. Rapid epidemic expansion of the SARS-CoV-2 Omicron variant in southern Africa. Nature. 2022;603(7902):679–686. doi:10.1038/s41586-022-04411-y

22. Wang Y, Zhang L, Li Q, et al. Development and validation of a multiplex RT-qPCR assay for simultaneous detection of SARS-CoV-2 and influenza A/B. J Clin Virol. 2021;144:104998. doi:10.1016/j.jcv.2021.104998

23. Itokawa K, Sekizuka T, Hashino M, et al. Disentangling primer interactions improves SARS-CoV-2 genome sequencing by multiplex tiling PCR. PLoS One. 2020;15(9):e0239403. doi:10.1371/journal.pone.0239403

24. Thermo Fisher Scientific. TaqPath COVID-19 CE-IVD RT-PCR Kit. Instructions for Use. Thermo Fisher Scientific; 2020.

25. FDA. SARS-CoV-2 Viral Mutations: Impact on COVID-19 Tests. Silver Spring, MD: US Food and Drug Administration; 2023.

26. Wolf JM, Wolf LM, Bello GL, et al. Evaluation of the performance of SARS-CoV-2 RT-qPCR commercial kits on detecting the E484K mutation. J Virol Methods. 2021;298:114283. doi:10.1016/j.jviromet.2021.114283

27. Lorenz R, Bernhart SH, Höner zu Siederdissen C, et al. ViennaRNA Package 2.0. Algorithms Mol Biol. 2011;6:26. doi:10.1186/1748-7188-6-26

28. Reusken CBEM, Broberg EK, Haagmans B, et al. Laboratory readiness and response for SARS-CoV-2 variants. Euro Surveill. 2021;26(14):2100349. doi:10.2807/1560-7917.ES.2021.26.14.2100349

29. Sanderson T, Hisner R, Donovan-Banfield I, et al. A molnupiravir-associated mutational signature in global SARS-CoV-2 genomes. Nature. 2023;623(7987):594–600. doi:10.1038/s41586-023-06649-6

30. Hill SC, Geoghegan JL, Holmes EC. Estimating the relative fitness of SARS-CoV-2 variants. Nat Rev Genet. 2022;23(8):464–477. doi:10.1038/s41576-022-00483-y

31. Grubaugh ND, Hodcroft EB, Fauver JR, et al. Public health actions to control new SARS-CoV-2 variants. Cell. 2021;184(5):1127–1132. doi:10.1016/j.cell.2021.01.035

32. Yang J, Noyce RS, Phelps NB, et al. Development of a multiplex PCR assay for detection of five emerging pathogens. J Clin Microbiol. 2022;60(1):e01736–21. doi:10.1128/JCM.01736-21

33. Stoddard CI, Galloway J, Chu HY, et al. Development and validation of a multiplex RT-PCR assay for SARS-CoV-2. PLoS One. 2021;16(5):e0251504. doi:10.1371/journal.pone.0251504

34. Lo SW, Jamrozy D, Chau KK, et al. Evaluation of a multiplex PCR assay for the identification of respiratory pathogens. J Med Microbiol. 2020;69(5):736–743. doi:10.1099/jmm.0.001191

35. World Health Organization. *Genomic sequencing of SARS-CoV-2: a guide to implementation for maximum impact on public health*. Geneva: WHO; 2021. Disponible en: https://www.who.int/publications/i/item/9789240018440

36. Peacock TP, Brown JC, Zhou J, et al. The SARS-CoV-2 variant, Omicron, shows rapid replication in human primary nasal epithelial cultures. Nature. 2022;602(7898):714–720. doi:10.1038/s41586-021-04352-y

37. Robishaw JD, Alter SM, Solano JJ, et al. Genomic surveillance to combat COVID-19: challenges and opportunities. Lancet Microbe. 2021;2(9):e423–e424. doi:10.1016/S2666-5247(21)00171-6

38. Bordoy AE, Saludes V, Panisello Yagüe D, et al. Monitoring SARS-CoV-2 variant transitions using differences in diagnostic cycle threshold values of target genes. Sci Rep. 2022;12(1):21818.

39. Bustin SA, Nolan T. Pitfalls of quantitative real-time reverse-transcription polymerase chain reaction. J Biomol Tech. 2004;15(3):155–166.

40. Thermo Fisher Scientific. TaqMan SNP Genotyping Assays User Guide. Publication No. 4332856 Rev. C. 2020.

41. Kandeel M, Mohamed MEM, Abd El-Lateef HM, Venugopala KN, El-Beltagi HS. Omicron SARS-CoV-2 variant: a mutational perspective. Int J Mol Sci. 2022;23(24):16153. doi: 10.3390/ijms232416153.

42. Outbreak.info. Variant: BA.1 [Internet]. La Jolla (CA): Scripps Research; 2025 [citado 4 de junio de 2026]. Disponible en: https://outbreak.info/situation-reports/variants/ba.1.

43. Chen J, Malone B, Llewellyn E, Grasso M, Shelton PMM, Olmeda-Martínez M, et al. Mutations in SARS-CoV-2 variant nsp6 enhance type-I interferon antagonism. Emerg Microbes Infect. 2023;12(1):2209208. doi: 10.1080/22221751.2023.2209208.

